# Perception and satisfaction of pharmacists’ roles and services provided in University of Nigeria Nsukka

**DOI:** 10.1101/2023.05.28.23290645

**Authors:** Judith Chinaecherem Azor, Adaobi Uchenna Mosanya, Blessing Onyinye Ukoha-Kalu

## Abstract

**Background:** Pharmacists are drug specialists in the society. The roles of pharmacists have extended beyond the typical product-oriented duties of dispensing, delivering medication and medical supplies to more patient-centered care. Patient satisfaction is a key indicator for healthcare quality and a metric to identify aspects that need improvement.

**Objective:** The aim of this study was to evaluate the public perception of pharmacists’ roles and satisfaction with the services they provide.

**Method:** Using a self-administered questionnaire, a cross-sectional descriptive study was conducted, data were analyzed using Statistical Package for Social Sciences (SPSS) version 25. Out of the 600 distributed questionnaire, 592 completed questionnaires were retrieved.

**Results:** Majority of the respondents were between the ages 18 and 30 years (88.5%) and had secondary school education as their highest level of educational qualification (73.6%). Higher proportion of the female respondents had a positive perception (72.4%). Also, they had higher satisfaction from the services (72.5%). Educational qualification (p=0.001), gender (p= 0.027), age (p= 0.006) and employment (p< 0.001) were significantly associated with the level of satisfaction from the services provided by pharmacists.

**Conclusion:** A good proportion of the members of the University community had a positive perception of the duties of pharmacists and were moderately satisfied with the services they provide. Steps should be taken to increase the amount and quality of time pharmacists spend with each patient.

## INTRODUCTION

Pharmacy profession is crucial to the healthcare system [1]. According to the American Pharmacists Association (APhA), pharmacists are health specialists who help patients take their prescriptions as effective as possible [2]. As members of the therapeutic team who are directly involved in patient care as well as dispensing pharmaceuticals, pharmacists play a crucial part in the healthcare system [3]. The roles of pharmacists are extending beyond the typical product-oriented duties of dispensing medicines and medical supplies to a more patient-centered care. Pharmacists are leaving factionalism behind and patient centered pharmaceutical care has become the mainstay of practice [4]. The maiden description of the concept was published in 1990 by Hepler and Strand as Pharmaceutical care. This is a responsible provision of drug therapy for the purpose of achieving definite outcomes that improve a patient’s quality of life [4]. Pharmaceutical care is the practice in which the drug related needs of a patient is taken care of by the healthcare professional and he/ she is held accountable for this commitment [5]. The main goal of pharmaceutical care is to improve patient outcomes; hence, it is outcome oriented. Studies have shown that provision of pharmaceutical care in hospitals had great value in clinical and economic outcomes[6]. Patients satisfaction is one of the humanistic outcomes derived from pharmaceutical care which has been used as a tool in assessing the quality of health care.

Patient’s perception affects awareness and attitude of the patient towards effectiveness and safety of drugs, acceptability of medication and patient adherence [7]. Patient’s perception of pharmacists is seen in the level of trust and confidence that patients have in the pharmacists, attitudes towards them and their expectations. This perception is affected by numerous social and health related factors [3]. As pharmacy practice moves from being product oriented, pharmacists are no longer just “dispensers of medication”, the need for better pharmacist-patient relationship increases, as this relationship is thought to influence patient health outcomes. A good patient-pharmacist relationship has the pharmacist and the patient working together and can lead to better patient satisfaction and compliance with the treatment plan.

An important measure employed to know the quality of healthcare, identifying potential areas for improvement, in order to increase the effectiveness of the healthcare system is patient satisfaction [8]. Understanding public’s need, expectation and satisfaction would help in the progress of pharmacy services in any country and also assist pharmacists, organizations and regulatory agencies that rely on consumer’s views, opinions and needs to modify the quality of services provided [9]. Satisfied customers are more likely to build trusting relationships with the personnel providing the healthcare services, bringing about higher cooperation levels and better health outcomes [10]. A key indicator to comparing the quality of services in various patient care services, systems and programs is Patient Satisfaction and this indicator is useful for providing better healthcare services and ensuring greater patient compliance [11]. Patients’ relationships with pharmacists and their expectations surrounding the services they provide influence the role and social standing of pharmacists [3].

There is no existing published literature on the public perception of pharmacist roles among the inhabitants of the university of Nigeria, Nsukka campus, a semi-urban community in south-east Nigeria.

## METHOD

This study is reported following the Strengthening the Reporting of Observational Studies in Epidemiology (STROBE) guidelines (see supplementary material)

### Study Design

This prospective cross-sectional survey, was conducted within members of the university community from February to June 2022. A questionnaire was developed for use as an exit survey for patients who came to the hospital and were in the five out-patient pharmacy units for either medication refill, get medicine information or collect medicine.

### Study Population

The study was conducted in the University of Nigeria, a federal government-owned university in south-east Nigeria. The study population consisted of members of the university community aged 18 years and above, students, and members of staff. An estimated population of 30,000 was used for this study [12]. University community members who failed to give consent were excluded. Similarly, pharmacists, pharmacy students or professionals in any health or medical field were not eligible for the study.

### Sample Size Determination/ Sampling Technique

Sample size was extrapolated using the Raosoft sample set calculator [13]. To calculate sample size using the Raosoft calculator, margin of error that can be accepted was 5%, confidence level was 95%, population size used was 30,000 and response distribution was 50%. According to the criteria above and the sample population, the sample size for the study was 600. A convenience sampling system was adopted for this study in the selection of respondents. The respondents were asked to fill the questionnaires immediately and completed ones were retrieved immediately.

### Study Procedure and Study Instrument

A validated questionnaire from the study of Jose *et al*. in Oman [9] was adapted, with some modifications. The questionnaire comprised of close ended questions in three sections: demographics, perception related and satisfaction related sections. Response to the questions was based on a five-point Likert scale. Samples of the self-administered questionnaires were distributed to three lecturers of the department of clinical pharmacy for face and content validation. Collection of data was done using convenience sampling method.

### Data Analysis

Completed questionnaires were collected, coded and analyzed using the SPSS version 25. Descriptive statistics was used to summarize data, scoring of responses was done for all items as the mean score and standard deviations according to original 5-point Likert scale. Inferential statistics such as Pearson’s Chi square test was used to evaluate the relationship between participants’ socio-demographic characteristics and satisfaction.

### Ethical Considerations

Ethical approval was obtained from the Faculty Research Ethics Committee, Faculty of Pharmaceutical Sciences, University of Nigeria, Nsukka (Ethical approval number: FPSRA/UNN/22/0045). In order to ensure that the participants gave informed consent, we explained the aim of the study by ensuring that they understood its contents. Also they were made to understand that participation was voluntary. We certify that all procedures followed the rules outlined in the Declaration of Helsinki.

## RESULTS AND DISCUSSIONS

From the study, 592 respondents took part in the survey. Table 1 below shows the socio-demographic characteristics of the respondents. Majority of the respondents were students (77.7%) between the ages 18-30 (88.5%) while a greater percentage of them had only secondary school education (73.6%). A good proportion of the respondents (77.2%) had visited the pharmacy less than 10 times in the last one year.

**Table 1:** Socio-demographic characteristics of study participants

|  |  | Frequency | Percentage |
| --- | --- | --- | --- |
| **Age(years)** | 18-30 | 524 | 88.5 |
|  | 31-45 | 42 | 7.1 |
|  | 46-60 | 22 | 3.7 |
|  | Above 60 | 4 | 0.7 |
| **Gender** | Males | 185 | 31.3 |
|  | Females | 407 | 68.8 |
| **Educational qualification** | No formal Education | 10 | 1.7 |
|  | Primary school | 5 | 0.8 |
|  | Secondary school | 436 | 73.6 |
|  | Higher education | 141 | 23.8 |
| **Employment** | Employee | 69 | 11.7 |
|  | Self- employed | 49 | 8.3 |
|  | Unemployed | 14 | 2.4 |
|  | Student | 460 | 77.7 |
| **Number of visits** | Less than 10 times | 457 | 77.2 |
|  | 10- 19 | 111 | 18.8 |
|  | 20- 30 | 18 | 3.0 |
|  | More than 30 times | 6 | 1.0 |

Table 2 shows the results of one-sample t test of the perception and satisfaction scores of the respondents, the item “ Pharmacists should check my prescriptions for accuracy…” had the highest positive perception with a mean score of 4.59 (standard deviation 0.697). The statement “ Pharmacists are mere vendors” had the least score with a mean of 3.15 (±1.207). A low perception score among the respondents indicates that Pharmacists are regarded not as mere vendors only. Many of the respondents agreed that pharmacists should let them know how to use their medications (4.55 ±0.694). A good number of respondents also agreed that pharmacists should answer their drug related questions (4.48± 0.719).

**Table 2:** Statistics for respondents’ perception scores (N = 592)

|  | Mean | Standard<br>Deviation | 95% confidence level<br>Lower | Upper | t |
| --- | --- | --- | --- | --- | --- |
| Drug matter | 4.46 | 0.704 | 4.40 | 4.51 | 154.021 |
| Minor ailments | 4.06 | 0.844 | 3.99 | 4.12 | 116.896 |
| Mere vendors | 3.15 | 1.207 | 3.06 | 3.25 | 63.573 |
| Integral part of healthcare system | 4.34 | 0.790 | 4.28 | 4.40 | 133.668 |
| Provide extended services | 4.09 | 0.883 | 4.02 | 4.16 | 112.699 |
| Dispensers of drugs | 4.29 | 0.796 | 4.20 | 4.33 | 130.247 |
| Check my prescriptions for accuracy | 4.59 | 0.697 | 4.53 | 4.65 | 160.282 |
| Let me know how to use medication | 4.55 | 0.694 | 4.50 | 4.61 | 159.767 |
| Answer my drug related questions | 4.48 | 0.719 | 4.42 | 4.54 | 151.537 |
| Information on the use of medicines | 4.32 | 0.797 | 4.26 | 4.39 | 131.979 |
| Advice patients on general health issues other than about drugs | 3.88 | 1.061 | 3.80 | 3.97 | 89.079 |
| Information on drug related matters | 4.00 | 0.871 | 3.93 | 4.07 | 111.878 |
| Questions asked by my pharmacists | 3.96 | 0.929 | 3.89 | 4.04 | 103.813 |
| Privacy maintained by the pharmacist | 3.92 | 1.006 | 3.84 | 4.01 | 94.948 |
| Level of knowledge | 4.08 | 0.897 | 4.00 | 4.15 | 110.555 |
| Language used by pharmacist | 3.88 | 0.937 | 3.80 | 3.95 | 100.635 |
| Response pharmacists provide | 3.90 | 0.904 | 3.83 | 3.97 | 104.961 |
| Time spent by my pharmacist | 3.54 | 1.015 | 3.46 | 3.62 | 84.815 |
| Relationship pharmacist maintains | 3.64 | 1.040 | 3.55 | 3.72 | 85.105 |
| Kind of information the pharmacists provide | 3.89 | 0.996 | 3.81 | 3.97 | 95.114 |

Figure 1 shows the perception of some selected pharmacists roles and satisfaction with a couple of pharmaceutical services. More than 50% of the respondents either were on the fence or agreed that pharmacists are mere vendors of drugs. On the other hand, 64% of them perceived the role of pharmacists in health promotion. About 70% of them were satisfied with the privacy that the pharmacists maintain while offering pharmaceutical care services. More than a quarter of the respondents were neither satisfied nor dissatisfied with the time that pharmacists spend with them individually.

**Figure 1.**
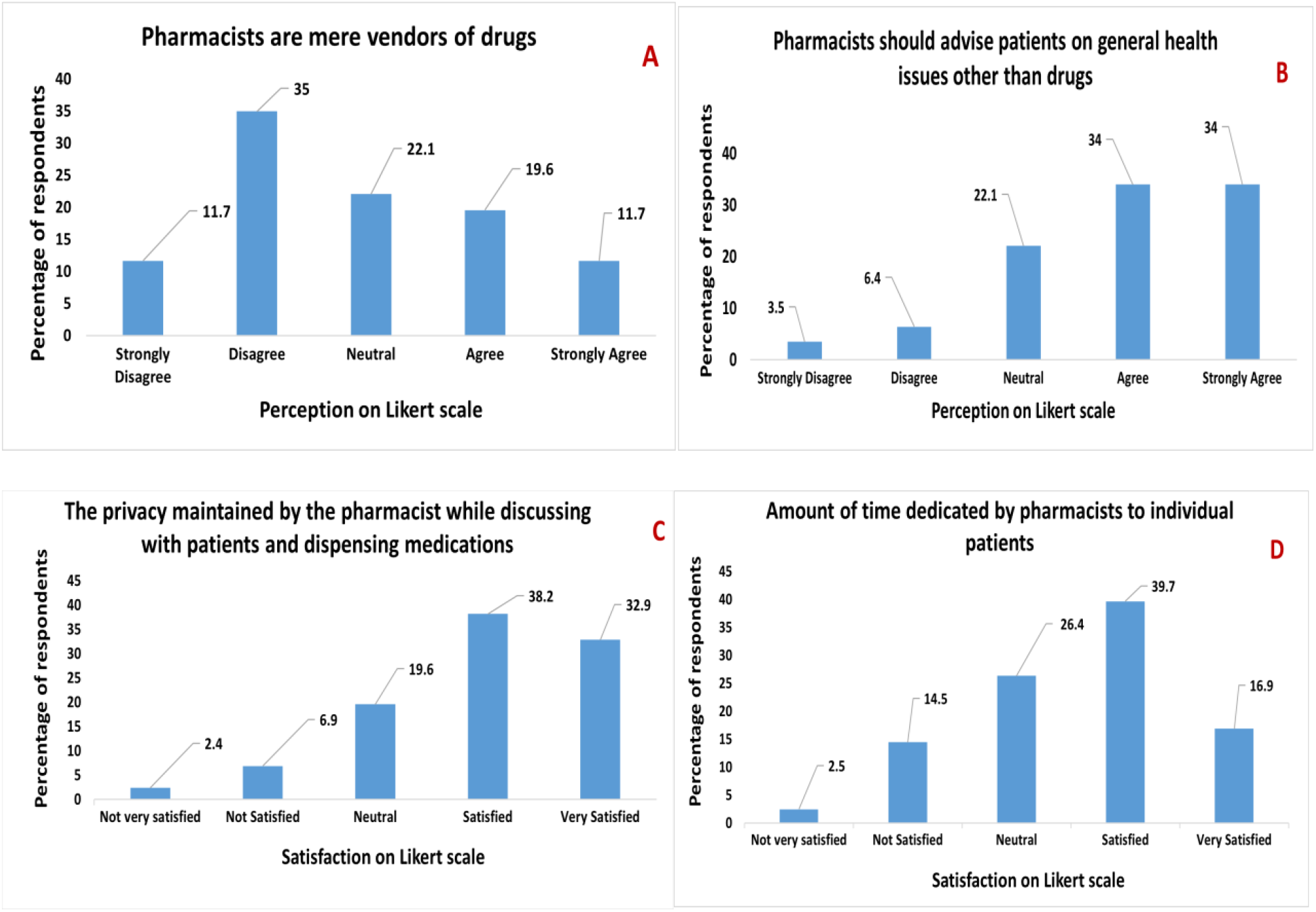
Public perception of pharmacists as mere vendors of drugs (A), public perception of pharmacists role in health promotion (B), public satisfaction with the privacy maintained by pharmacists while offering pharmaceutical care (C) and public satisfaction with the amount of time pharmacists spend with them individually (D).

Table 3 shows the Chi-square association of perception and satisfaction with socio-demographics. Majority of the respondents were between the ages 18-30 years (88.5%) has the best perception levels (89.4%) and highest satisfaction levels (92.5%).

**TABLE 3:** Association of perception and satisfaction with sociodemographic

|  |  | Total sample | Good perception (%) | P-value | High satisfaction | P-value |
| --- | --- | --- | --- | --- | --- | --- |
| **Age(years)** | 18-30 | 524 (88.5) | 269(89.4) | 0.622 | 302(92.5) | **.006** |
|  | 31-45 | 42 (7.1) | 19(6.3) |  | 16(4.8) |  |
|  | 46-60 | 22 (3.7) | 10(3.3) |  | 8(2.4) |  |
|  | Above 60 | 4 (0.7) | 3(1.0) |  | 1(0.3) |  |
| **Gender** | Males | 185(31.3) | 83(27.6) | **0.050** | 92(27.5) | **.027** |
|  | Females | 407(68.8) | 218(72.4) |  | 242(72.5) |  |
| **Educational qualification** | No formal education | 10(1.7) | 5(1.7) | 0.125 | 4(1.2) | **.001** |
|  | Primary school | 5(0.8) | 1(0.3) |  | 1(0.3) |  |
|  | Secondary school | 436(73.6) | 233(74.4) |  | 266(79.6) |  |
|  | Higher education | 141(23.8) | 62(20.6) |  | 63(18.9) |  |
| **Employment** | Employee | 69(11.7) | 31(10.3) | 0.114 | 26(7.8) | <<br>**0.001** |
|  | Self employed | 49(8.3) | 18(6.0) |  | 18(5.4) |  |
|  | Unemployed | 14(2.4) | 7(2.3) |  | 8(2.4) |  |
|  | Student | 460(77.7) | 245(81.4) |  | 282(84.4) |  |
| **Number of visits to the pharmacy in the last year** | Less than 10 times | 457(77.2) | 228(75.7) | 0.762 | 261(78.1) | .899 |
|  | 10-19 | 111(18.8) | 59(19.6) |  | 61(18.3) |  |
|  | 20-30 | 18(3.0) | 11(3.7) |  | 9(2.7) |  |
|  | More than 30 times | 6(1.0) | 3(1.0) |  | 3(0.9) |  |

Although, it should be noted that majority of the respondents who participated in the study were students of the University community. Females had positive perception (72.4%) and higher satisfaction (72.5%) compared to the males (satisfaction: 27.5%, perception: 27.6%). However, this finding was not significant for the level of perception and was significant for the level of satisfaction, hence, it can be said that gender was not significantly associated with the level of perception but was significantly associated with the level of satisfaction. As seen in this study, respondents with secondary school and those with higher education as their highest levels of educational qualification had positive perception and high level of satisfaction compared to those with no formal education. The level of educational qualification was not significantly associated with the level of perception of the roles of pharmacists but was significantly associated with the level of satisfaction of the services provided.

Number of visits to the pharmacy was neither significantly associated with the level of perception nor with the level of satisfaction. This research showed that respondents who visited the pharmacy less than ten times in the last year, had positive perception of the roles of pharmacists and were highly satisfied with the services provided.

This study showed that many respondents agreed that pharmacists should be provide clients with advice on general health issues, apart from drugs. In this era of technology, where information can be easily accessed on the internet [9], it is good to see that many respondents are of the opinion that Pharmacists should educate them on other health issues. Many participants were satisfied with the quality and amount of information gotten from the pharmacists on drug related matters. However, they were not satisfied with the amount of time spent by the pharmacist with each patient. The level of satisfaction was moderate with respect to the relationship the pharmacists try to maintain with the patients. Patient follow-up is an important aspect of pharmaceutical care, as it helps to monitor patient adherence to medication and ensure that drug therapy problems are prevented.

Patient-pharmacist relationship based on caring, trust, cooperation and mutual decision making helps to obtain better patient outcomes and effective drug therapy results. Patient satisfaction has been widely used to assess the quality of healthcare by identifying potential areas for improvement [8]. Many participants were satisfied with the type and amount of quality information accorded them by the pharmacists on drug related matters. However, they were not satisfied with the amount of time spent interacting with the pharmacists. These findings are similar to the results of the study carried out in Dubai by Ali *et al*. where they reported that the rightness of therapy, including drug information and drug interactions usually required additional time to be consumed by the patient [14]. Therefore, the quality of information and communication on drug supplied is dependent on the time provided for each patient by the pharmacist. This shows the relationship between time provided by pharmacist and the level of satisfaction.

Many of the respondents were within the ages 18-30 years, these respondents had positive perception of the duties of pharmacists and the level of satisfaction with the services was high. Result of this study is similar to that conducted by Aniza *et al*., which showed that increase in age significantly correlated with lower level of patient satisfaction [8]. Research by Rensburg *et al*. found that older patients are more vulnerable and have higher expectations when it comes to meeting their medicine related needs, due to chronic diseases [15]. Greater expectations from the quality of healthcare services provided may be seen among those with higher education [8]. As seen in this study, respondents with secondary school and those with higher education as their highest levels of educational qualification had positive perception and high level of satisfaction compared to those with no formal education. Similar study in Korea showed that patients having higher education had lower satisfaction compared with patient lower education [16]

This research showed that respondents who visited the pharmacy less than ten times in the previous year, had positive perception of the duties of pharmacists and were highly satisfied with the services accorded them. This was different from the result obtained by Aniza *et al*., which showed that higher number of visits to the pharmacy was significantly associated with higher patient satisfaction [8], the reason could be that patient involved in the study were on medication for chronic diseases, which requires them to visit the pharmacy more often.

## CONCLUSION

This study indicated that majority of the members of the University community had a positive perception of the roles of pharmacists and are moderately satisfied with the services provided to them by pharmacists. Employment, gender, age and educational qualification were significantly associated with the level of satisfaction of the services provided. Pharmaceutical care services provided at the time of dispensing are a crucial component of optimizing prescription therapy. Steps should be taken to improve the amount of time the pharmacists spend with each patient and the relationship between the pharmacist and the patient be cordial, thereby improving patient satisfaction. Furthermore, there is need to identify gaps in the public’s understanding and awareness of the role of the pharmacists.

## Strengths and Limitations of the Study

Our findings is one of the first studies to look at the public perception of pharmacists roles and services in a semi-urban community in Nigeria. This will contribute to the growing body of evidence that supports how pharmacists can build relationship with their patients and how this can improve patient outcomes.

A limitation of the study is inability to make generalization because the sample from the University community in south east Nigeria may not be broadly representative with respective to the services provided by pharmacists in other geopolitical zones in Nigeria.

## Research and Clinical Implications

- Qualitative interviews with pharmacists in the hospital and community settings could be considered in future studies to see how to improve the quality of healthcare services provided, as well as patients’ experiences.
- Pharmacists, in the hospital and community settings should focus on improving communication with patients, providing quality drug information to improve patients’ experiences.
- Also strategic awareness programmes should be implemented to improve the public perception of pharmacists role.

## Data Availability

All data produced in the present study are available upon reasonable request to the authors

## Authors’ Contributions

JCA, AUM and BOU-K devised the study and developed/refined the main conceptual ideas. JCA and BOU-K led the study protocol development, ethical application, and gaining approval, with input from the whole team. JCA and AKA undertook recruitment and data collection. AUM and BOU-K provided support for study conduct, data collection and analyses. JCA, AKA and BOU-K drafted the manuscript. All authors helped refine and re-draft the manuscript and approved the final version.

## Acknowledgements

The authors thank all the participants for their cooperation in data collection and in the preparation of this manuscript.

## Conflict of interest

The authors have no conflict of interest to declare.

## Funding

This research received no specific grant from any funding agency in the public, commercial, or not-for-profit sectors.

## Availability of data and materials

The dataset(s) supporting the conclusions of this article are included within the article (and its additional file(s)).

## Notes

### Competing Interest Statement

The authors have declared no competing interest.

### Funding Statement

This study did not receive any funding

### Author Declarations

Ethical approval was obtained from the Faculty Research Ethics Committee, Faculty of Pharmaceutical Sciences, University of Nigeria, Nsukka (Ethical approval number: FPSRA/UNN/22/0045).

